# Oropharyngeal Microbiome Profiled at Admission is Predictive of the Need for Respiratory Support Among COVID-19 Patients

**DOI:** 10.1101/2022.02.28.22271627

**Authors:** Evan S Bradley, Abigail L. Zeamer, Vanni Bucci, Lindsey Cincotta, Marie-Claire Salive, Protiva Dutta, Shafik Mutaawe, Otuwe Anya, Christopher Tocci, Ann Moormann, Doyle V. Ward, Beth A. McCormick, John P Haran

## Abstract

The clinical course of infection due to respiratory viruses such as Severe Acute Respiratory Syndrome Coronavirus 2 (SARS-CoV2), the causative agent of Coronavirus Disease 2019 (COVID-19) is thought to be influenced by the community of organisms that colonizes the upper respiratory tract, the oropharyngeal microbiome. In this study, we examined the oropharyngeal microbiome of suspected COVID-19 patients presenting to the Emergency Department and an inpatient COVID-19 unit with symptoms of acute COVID-19. Of 115 enrolled patients, 74 were confirmed COVID-19+ and 50 had symptom duration of 14 days or less; 38 acute COVID-19+ patients (76%) went on to require respiratory support. Although no microbiome features were found to be significantly different between COVID-19+ and COVID-19-patients, when we conducted random forest classification modeling (RFC) to predict the need of respiratory support for the COVID-19+ patients our analysis identified a subset of organisms and metabolic pathways whose relative abundance, when combined with clinical factors (such as age and Body Mass Index), was highly predictive of the need for respiratory support (F1 score 0.857). Microbiome Multivariable Association with Linear Models (MaAsLin2) analysis was then applied to the features identified as predicative of the need for respiratory support by the RFC. This analysis revealed reduced abundance of *Prevotella salivae* and metabolic pathways associated with lipopolysaccharide and mycolic acid biosynthesis to be the strongest predictors of patients requiring respiratory support. These findings suggest that composition of the oropharyngeal microbiome in COVID-19 may play a role in determining who will suffer from severe disease manifestations.

**Importance:** The microbial community that colonizes the upper airway, the oropharyngeal microbiome, has the potential to affect how patients respond to respiratory viruses such as SARS-CoV2, the causative agent of COVID-19. In this study, we investigated the oropharyngeal microbiome of COVID-19 patients using high throughput DNA sequencing performed on oral swabs. We combined patient characteristics available at intake such as medical comorbidities and age, with measured abundance of bacterial species and metabolic pathways and then trained a machine learning model to determine what features are predicative of patients needing respiratory support in the form of supplemental oxygen or mechanical ventilation. We found that decreased abundance of some bacterial species and increased abundance of pathways associated bacterial products biosynthesis was highly predictive of needing respiratory support. This suggests that the oropharyngeal microbiome affects disease course in COVID-19 and could be targeted for diagnostic purposes to determine who may need oxygen, or therapeutic purposes such as probiotics to prevent severe COVID-19 disease manifestations.

## Introduction

Coronavirus Associated Infectious Disease 2019 (COVID-19) is caused by infection with the severe acute respiratory syndrome coronavirus 2 (SARS-CoV2). COVID-19 has sickened nearly 50 million and caused in excess of 770,000 deaths in the United States alone^1^. Some individuals develop severe disease and death while others present with only mild or no symptoms^2^. There are known clinical factors that are associated with risk of severe disease such as age, diabetes, high blood pressure, and obesity^3^, but predicting whether an individual patient will require hospitalization or respiratory support, or can recover safely at home has important implications for healthcare resource utilization. Currently, clinical factors such as age, BMI, and medical comorbidities, in combination with initial vital sign measurements, need for oxygen support, and clinical laboratory testing, are used to predict clinical decompensation and the need for ICU level of care--even the best algorithms, however perform only with an accuracy of 70-80%^4,5^. There are likely other individual factors that determine how a patient responds to COVID-19 and may play a role in determining disease manifestations, such as the need for respiratory support^6^.

The oropharyngeal and nasopharyngeal microbiomes, the collection of organisms that colonize the human upper airway, have been hypothesized to influence the host immune responses to respiratory viral and bacterial infections^7^. Commensal bacterial species of the nasopharynx can modulate the immune response to influenza virus infection in a potentially protective way^8,9^. Conversely, viral co-infection in the upper airway and lungs may promote bacterial pathogens by liberating nutrients or exposing adhesion molecules^10,11^ leading to more severe disease and secondary bacterial infection. Here we hypothesize that information from the oropharyngeal microbiome along with clinical variables routinely collected at admission are predictive of the clinical trajectory of COVID-19 cases and specifically of the need of receiving respiratory support. To test this hypothesis we investigated the oropharyngeal microbiome of individuals presenting with symptoms suggestive of COVID-19 and positive clinical testing for COVID-19. We used machine learning-based modeling to determine oropharyngeal microbiome signatures among COVID-19 patients examine associations between microbiome features patients going on to require respiratory support, and to quantify the ability of microbiome features to predict the need for respiratory support. We then inspect the determined microbiome-clinical outcome associations to possibly explain why some patients need respiratory support during a SARS-CoV2 infection.

## Results

### Patient Characteristics

Clinical data, demographic and comorbidity data are presented in Table 1. Our filtering and subject categorization scheme is shown in Figure 1. Our final analysis cohort consisted of 74 COVID-19+ patients. Of COVID-19+ cohort, 50 had known symptom duration of less than 14 days, of these 38 (76%) required some form of respiratory support. With the exception of Body Mass Index (BMI) (a.o.v p < 0.05) COVID-19+ patients requiring respiratory support, and those that did had similar characteristics. The overall mean age of the final cohort was 68 (SD 15.24), 50% were female, the majority of patients identified as Hispanic or Latino (76%) and white (64%). Within the acute COVID+ cohort (see Figure 1), 12 (24%) patients never required any respiratory support, 18 (36%) were treated with supplemental oxygen via nasal cannula, 3(6%) were treated with supplemental oxygen via facemask, 6 patients were treated positive pressure ventilation (12%), and 11 (22%) were intubated. There were 2 patients who died of COVID-19 but had Do Not Intubate (DNI) orders; accordingly, they were considered as having respiratory failure severe enough to be treated with intubation.

**Table 1.** Study Population Characteristic.

| Characteristic | Overall, N = 50<sup>1</sup> | Respiratory Support |  | p-value<sup>2</sup> |
| --- | --- | --- | --- | --- |
|  |  | no, N = 12<sup>1</sup> | yes, N = 38<sup>1</sup> |  |
| Age | 68.00 (15.24) | 60.83 (19.52) | 70.26 (13.12) | 0.15 |
| Caucasian | 32 / 50 (64%) | 5 / 12 (42%) | 27 / 38 (71%) | 0.089 |
| Black | 5 / 50 (10%) | 2 / 12 (17%) | 3 / 38 (7.9%) | 0.6 |
| Asian | 2 / 50 (4.0%) | 2 / 12 (17%) | 0 / 38 (0%) | 0.054 |
| Other | 11 / 50 (22%) | 3 / 12 (25%) | 8 / 38 (21%) | >0.9 |
| CCI | 4.50 (2.58) | 3.75 (3.05) | 4.74 (2.41) | 0.2 |
| hypertension | 33 / 50 (66%) | 7 / 12 (58%) | 26 / 38 (68%) | 0.7 |
| diabetes | 18 / 50 (36%) | 5 / 12 (42%) | 13 / 38 (34%) | 0.7 |
| asthma | 8 / 50 (16%) | 1 / 12 (8.3%) | 7 / 38 (18%) | 0.7 |
| COPD | 10 / 50 (20%) | 2 / 12 (17%) | 8 / 38 (21%) | >0.9 |
| OSA | 3 / 50 (6.0%) | 0 / 12 (0%) | 3 / 38 (7.9%) | >0.9 |
| Support type |  |  |  | <0.001 |
| None | 12 / 50 (24%) | 12 / 12 (100%) | 0 / 38 (0%) |  |
| Nasal cannula oxygen | 18 / 50 (36%) | 0 / 12 (0%) | 18 / 38 (47%) |  |
| Facemask/Oxymizer | 3 / 50 (6.0%) | 0 / 12 (0%) | 3 / 38 (7.9%) |  |
| NIPPV | 6 / 50 (12%) | 0 / 12 (0%) | 6 / 38 (16%) |  |
| Intubation | 11 / 50 (22%) | 0 / 12 (0%) | 11 / 38 (29%) |  |
| COVID Fatality | 8 / 50 (16%) | 0 / 12 (0%) | 8 / 38 (21%) | 0.2 |
| BMI | 29.12 (7.01) | 23.83 (5.11) | 30.79 (6.74) | 0.003 |
| male | 25 / 50 (50%) | 5 / 12 (42%) | 20 / 38 (53%) | 0.5 |
| Hispanic or Latino | 38 / 50 (76%) | 7 / 12 (58%) | 31 / 38 (82%) | 0.13 |
| Smoker, current | 1 / 50 (2.0%) | 1 / 12 (8.3%) | 0 / 38 (0%) | 0.2 |
| Smoker, former | 21 / 50 (42%) | 3 / 12 (25%) | 18 / 38 (47%) | 0.2 |
| shannon | 2.25 (0.62) | 2.50 (0.35) | 2.17 (0.66) | 0.2 |
| simpson | 0.80 (0.13) | 0.86 (0.04) | 0.78 (0.15) | 0.3 |
| invsimpson | 7.04 (3.69) | 7.70 (2.39) | 6.83 (4.02) | 0.3 |

<sup>1</sup> Mean (SD); n / N (%)
<sup>2</sup> Wilcoxon rank sum test; Fisher's exact test; Wilcoxon rank sum exact test; Pearson's Chi-squared test

**Figure 1.**
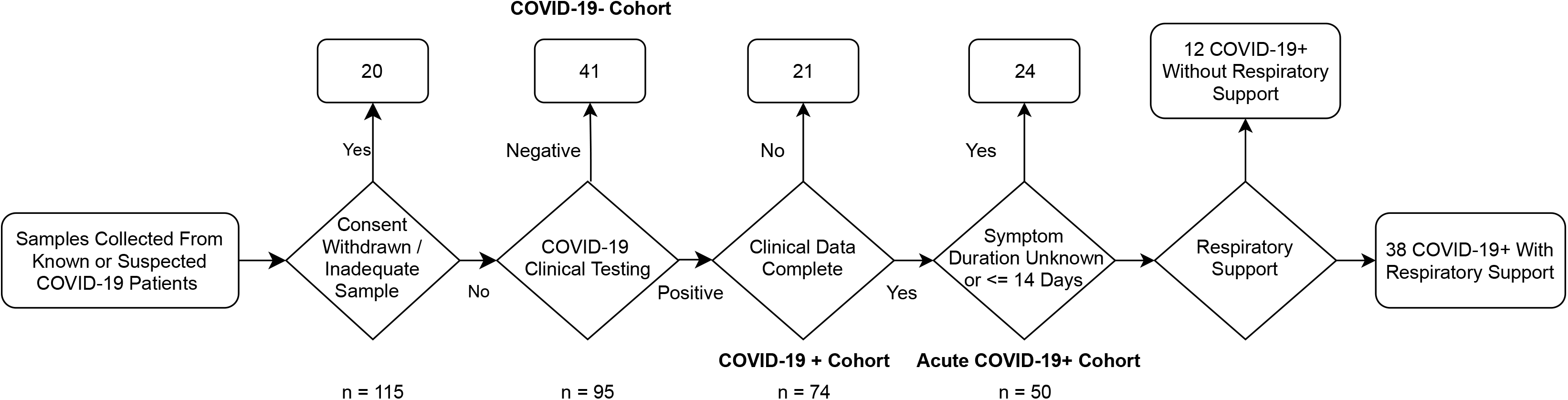
Study Enrollment Flow Chart.

### Features of the oropharyngeal microbiome are associated with need for respiratory support

We first directly compared abundances of microbiome features between COVID-19+ and COVID-19-patients utilizing the Wilcoxon Rank Sum test. When corrected for multiple comparisons, there were no bacterial species or metabolic pathway abundances that were significantly different between COVID-19+ and COVID-19-patients. We then trained RFC models to determine what clinical and microbiome features (species and metabolic pathway abundances) were predictive of need for respiratory support. We selected this model because previous work has demonstrated robust correlations between microbiome and clinical outcomes^12^. We chose this machine learning-based approach as it enables the use of non-normally distributed (species relative abundance) and a diverse set of variables (Shannon’s alpha diversity index, and numerical and categorical clinical covariates) as features in the same model thus allowing us to predict clinical response from complex multi-modal data^13^. To evaluate the performance of our models, we computed F1 score, the harmonic mean between precision and recall, which accounts for both prediction errors and the specific type of prediction error. Utilizing sample-level Shannon’s alpha diversity index and clinical covariates, which included age, BMI, race, ethnicity, selected medical comorbidities available at admission, the model performed well with a mean F1 score 0.857 ± 0.000 (Figure 2A). A model trained only on measured bacterial abundances performed comparably with a mean F1 score of 0.837 ± 0.005. A model including clinical covariates, select medical comorbidities, measured bacterial abundances, and sample-level Shannon’s alpha diversity index led to a similar predictive performance measured by a mean F1 score of 0.858 ± 0.009. These F1 scores indicate similar performance of clinical and microbial variables. Additional model statistics are included in Table S1. We examined the model that combined microbiome features and clinical covariates in more depth to compare directly how these factors were associated with the need for respiratory support.

**Figure 2.**
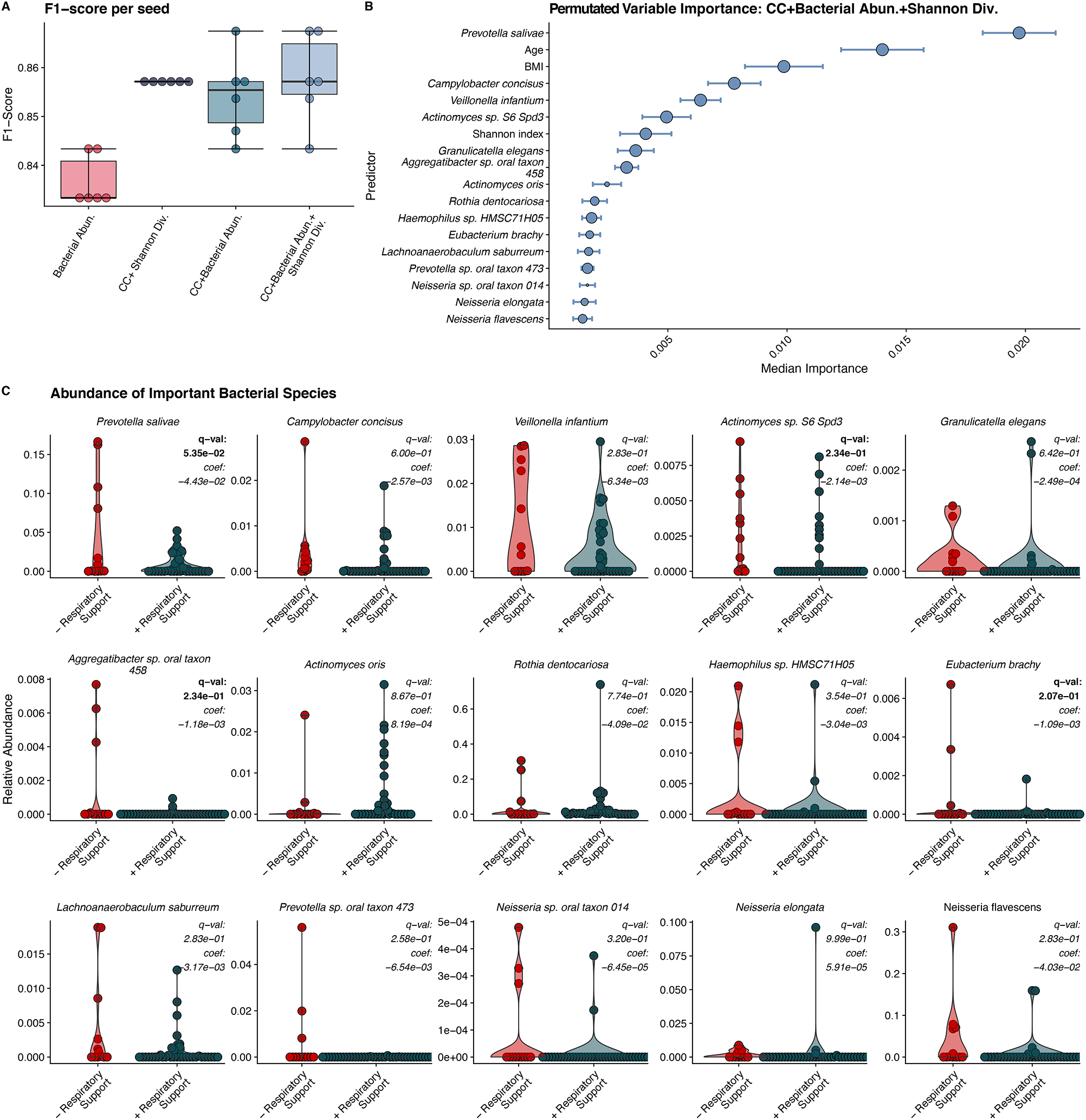
Results of Random Forest Classification Model. A) F1 scores of RFC models including clinical covariates (CC), individual bacterial abundances, and the combination of bacterial abundances, alpha diversity, and clinical covariates. All models perform well with models including microbiome data performing slightly better. B) Median ranked importance of model features including microbiome features and clinical data (median importance ± median absolute deviation). The size of the circle represents how often each feature was selected. The relative abundance of *Prevotella salivae* is the top predictor with the relative abundance of *Campylobacter concisus, Veillonela infantium* and *Actinomycetes* sp. S6-Spd3 and the Shannon diversity index also showing significant contributions. C. The relative abundance of the organisms determined to be important in predicting need for respiratory support by our RFC model. Q-values (BH adjusted p-values) and coefficients calculated via MaAslin2 are shown for each bug. By MaAsLin2, *Prevotella salivae, Eubacterium branchy, Actinomyces sp. S6 spd3 and, Aggregatibacter sp. oral taxon 45* were significantly associated (q < 0.25) with need for respiratory support and are bolded.

The aggregated permutated variable importance^14^ from the selected RFC model identified the relative abundance of *Prevotela salivae* as the most important predictor of the need for respiratory support (Figure 2B). Specifically, a decrease in *P. salivae* abundance was indicative of respiratory support need (Figure 2C). Notably, this organism is ranked higher than both patient age and BMI (Figure 2B), which are two clinical factors known to associate with severe COVID-19^3^. Other factors that were predictive of the need for respiratory support include decreases in Shannon’s alpha diversity and the decreases in the relative abundances of *Campylobacter concisus, Veillonella infantum*, and *Actinomycetes* species S6-Spd3 (Figure 2C).

To further explore connections between microbiome features and clinical covariates, we examined the association between the abundance of our 15 top-predicting microbes with clinical covariates using MaAsLin2. MaAsLin2 determines multivariable associations between clinical variables and microbiome data utilizing general linear models as opposed to a random forest^15^. This approach allows us to determine if specific microbiome predictors are associated with our clinical outcome of interest (need or O2 support) after explicitly controlling for the effect of possible confounding clinical covariates (i.e., age and BMI). Furthermore, MaAsLin2 analysis can also be considered an independent validation of our findings using a different methodology. The need for respiratory support was identified as significantly associated with four of the fifteen RFC-identified as important microbes, specifically, *P. salivae, Eubacterium branchy, Actinomyces sp. S6 spd3 and, Aggregatibacter sp. oral taxon 45* (Table 2). Age was found to be independently associated with abundance of *P. salivae*, and *Neisseria sp. oral taxon 014*. None of the top microbial predictors were found to associate with BMI. These results support the association between microbiome features and the need for respiratory support as these features were found to be significantly associated with this outcome utilizing an approach that specifically controls for potential confounders such as patients’ age and BMI.

**Table 2.** Results of MaAsLin Analysis on Bacterial Abundances.

| Clinical Covariate | Organism | Coefficient | Standard Error | p-value | q-value |
| --- | --- | --- | --- | --- | --- |
| Respiratory Support | *Prevotella salivae* | -0.044 | 0.013 | 0.0012 | 0.054 |
| Respiratory Support | *Eubacterium brachy* | -0.0011 | 0.0004 | 0.0092 | 0.21 |
| Age | *Prevotella salivae* | 0.00085 | 0.00034 | 0.015 | 0.22 |
| Respiratory Support | *Actinomyces sp S6 Spd3* | -0.0021 | 0.00094 | 0.028 | 0.23 |
| Respiratory Support | *Aggregatibacter sp oral taxon 458* | -0.0012 | 0.00053 | 0.031 | 0.23 |
| Age | *Neisseria sp oral taxon 014* | -2.28E-06 | 9.83E-07 | 0.025 | 0.23 |

**Table 3.**
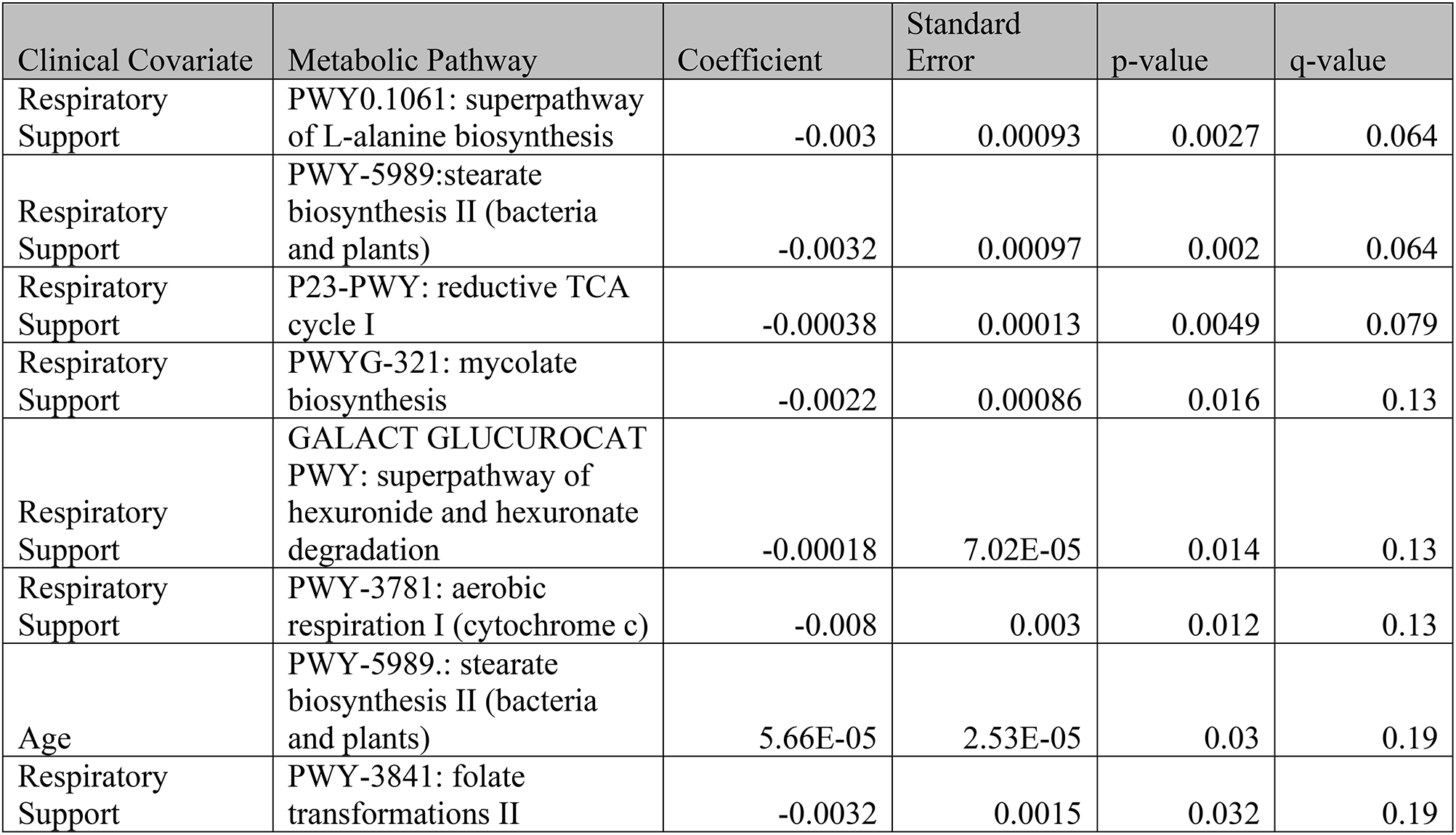
Results of MaAsLin Analysis on Metabolic Pathway Abundances.

Similar analysis was repeated on the samples profiled for the abundance of metabolic pathways using HUMAnN3^16^. The relative abundance of specific bacterial metabolic pathways was also highly predicative of the need for respiratory support (mean F1 score 0.804 ± 0.009) and adding clinical covariates available at admission to the model, resulted in a similar mean F1 score of 0.821 ± 0.004 (Figure 3A). Additional model statistics are included in Table S2. The metabolic pathways most important in predicting the need for respiratory are decreased abundance of LPS biosynthesis (CMP-3-D-*manno*-octulosonate and lipid IV A biosynthesis), mycolate biosynthesis, and trehalose degradation pathways and increased abundance of L-threonine, L-proline and inosine-5-phosphate pathways (Figure 3B,C). We examined the contribution of bacterial genera to two LPS biosynthetic pathways that were highly predicative of the need for respiratory support. We observed, less of the CMP-3-deoxy-D-*manno*-octulosonate pathway originating from *Prevotella* and large portion of this pathway is originating from *Pseudomonas* in patients who required respiratory support (Supplementary figure 1). A large contributor to the Lipid IVA biosynthesis pathway in patients who required respiratory support originated from *Aggrigatibacter*, a genus closely related to *Haemophilus influenzae*^*17*^. We similarly applied MaAsLin2 to the metabolic pathway predictors identified as important in our RFC. Seven of the top predictors identified also showed significant associations by MaAsLin2 with only one pathway (stearate biosynthesis) significantly associated with age as well. Notably, the relative abundance mycolic acid biosynthesis pathway was found to be a top predictor of the need for respiratory support and significantly associated with the need for respiratory support by MaAsLin2.

**Figure 3.**
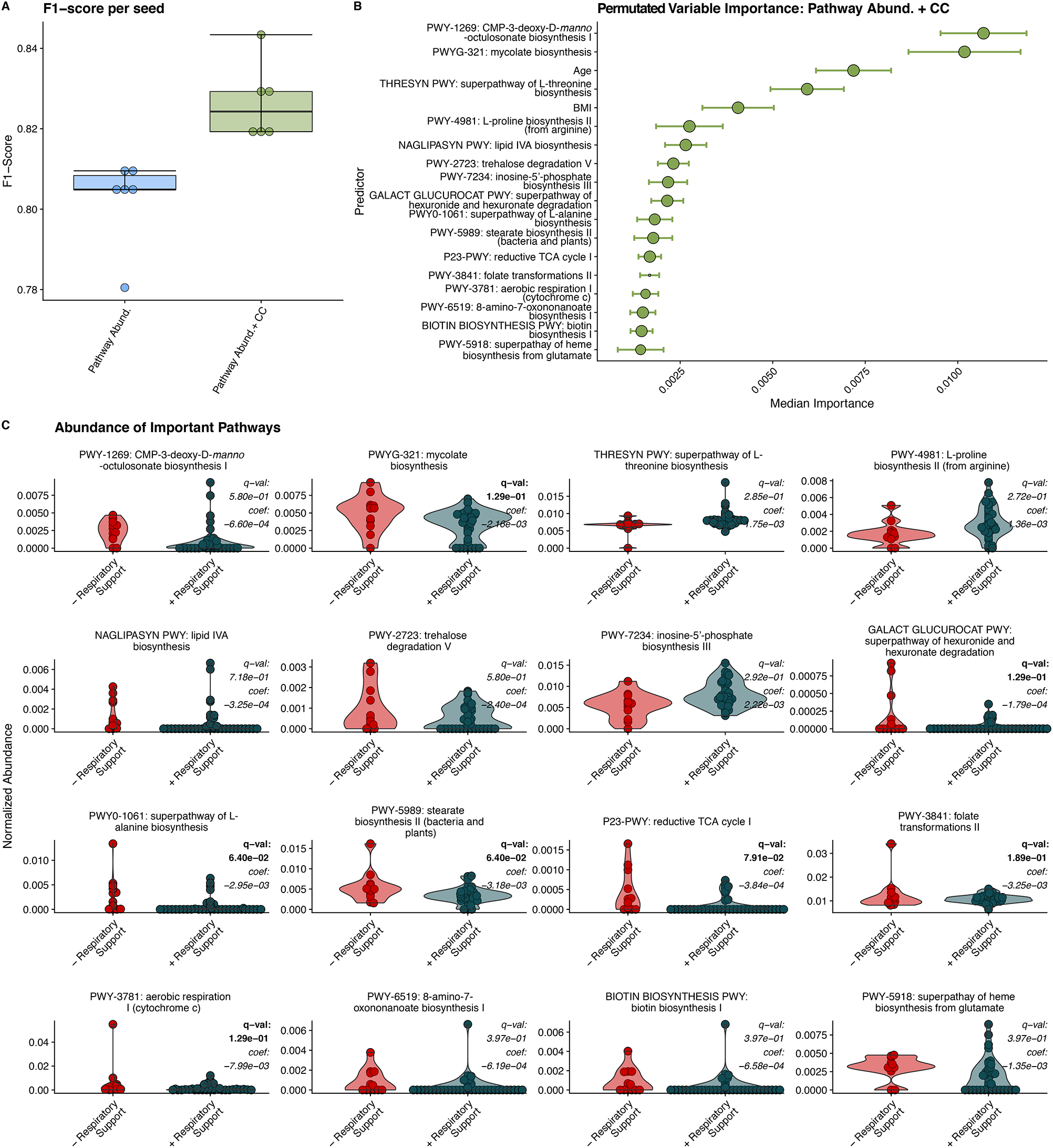
Random Forest Classification Using Metabolic Pathways. A) F1 scores of RFC models built on relative abundance of detected metabolic pathways and clinical covariates (CC). B) Median relative importance of variables in predicating the need for respiratory support within the trained with relative pathway abundances and clinical covariates (median importance ± median absolute deviation). C) Relative abundance of detected metabolic pathways in individuals requiring respiratory support and those not requiring respiratory support. MaAsLin2 derived q-values and coefficients are displayed for each pathway. Significant q values (q < 0.25) are bolded.

## Discussion

We show that the abundance of several Gram-negative and *Actinomyces* species and metabolic pathways associated with LPS, mycolic acid, and amino acid biosynthesis within the oropharyngeal microbiome are associated with COVID-19 patients developing the need for respiratory support and thus COVID-19 severity. The top predictors from our RFC predictive model were confirmed using an independent analysis based on generalized linear models. When examining important factors associated of the need for respiratory support, we found that decreased abundances of *P. salivae*, and an *Actinomyces* species were highly associated with the need for respiratory support in both analyses, suggesting the presence of these protective organisms is associated with COVID-19 patients not requiring respiratory support. A higher abundance of genes encoding the metabolic pathways for mycolate biosynthesis, L-alanine biosynthesis, stearate biosynthesis, folate transformation, and genes associated with aerobic utilization of hexuronides were identified in both analyses as associated with the need for respiratory support, with LPS biosynthesis genes (CMP-3-D-*manno*-octulosonate and lipid IV A biosynthesis) also found to be highly predictive in the RFC. These trends suggest that the most important microbiome factors in predicting the need for respiratory support are a higher abundance of some commonly detected oropharyngeal commensal bacteria and an increased abundance of pathways associated with bacterial product biosynthesis and aerobic respiration.

### *Prevotella* and LPS biosynthesis

Decreased *P. salivae* abundance was the strongest predictor of the need for respiratory in our RFC model and significantly associated with the outcome by MaAsLin2. Prior work has shown members of the *Prevotella* genus to be associated with COVID-19, with increased abundances of this genus as measured by 16S rRNA sequencing being associated with more severe disease^18^. This study included a similar number of COVID-19+ patients with similar disease severity but did not consider clinical variables when determining associations between organism abundance and disease severity, which we have included in our models. In addition, this was a study of nasopharyngeal swabs, as opposed to oral swabs, which is a distinctly different microbial community^7^ and may interact with SARS-CoV2 differently. *Prevotella* are Gram-negative anaerobic organisms and common oropharyngeal colonizers that have been implicated in periodontal disease^19^. Sequences encoding *Prevotella* house-keeping proteins such as the chaperonin GroEL and RNA polymerase were detected in metagenomic studies of the lungs of COVID-19 patients early in the outbreak^20^ and were hypothesized to play a role in the pathogenesis of COVID-19 lung disease^21^.

*Prevotella* has generally been implicated in chronic inflammation^22^ but is also part of the normal, healthy lung microbiome^23^. *P. salivae* has been shown in animal models to stimulate less inflammatory cytokine production and lead to less neutrophil chemotaxis than the Gram-negative respiratory pathogens *Morexella catarhallis* and *Haemophilus influenzae*^*24*^. It is hypothesized that a penta-acylated LPS produced by *Provetella*^25^ stimulates less innate-immune receptor activation than hexa-acylated LPS produced by Gram-negative respiratory pathogens and *Escherichia coli*^*22*^. This may represent an adaptation that allows *Prevotella* to colonize the upper airway without causing disease.

Our metagenomic analysis found that the abundance of two LPS biosynthetic pathways, CMP-3-deoxy-D-*manno*-octulosonate and lipid IV A biosynthesis, are the top predictors of the need for respiratory support in the RFC. CMP-3-deoxy-D-*manno*-octulosonate is a critical metabolite in LPS biosynthesis^26^, and lipid IVA is a precursor in the production of the lipid A core of LPS^27^. In our RFC model trained with metabolic pathways and clinical covariates, a higher abundance of these pathways appears protective, which initially seems counter-intuitive as LPS is known to generate substantial inflammation via the innate immune system activation^28^. When we examined the contribution of bacterial genera to the CMP-3-deoxy-D-*manno*-octulosonate biosynthesis pathway, we observed that less of the pathway originated from *Prevotella* in patients who required respiratory support and a larger portion of this pathway originates from *Pseudomonas*, a known respiratory pathogen capable of producing highly inflammatory LPS^29^. A large contributor to the Lipid IVA biosynthesis pathway originated from *Aggrigatibacter*, a genus closely related to *Haemophilus influenzae*^*17*^, which also produces highly inflammatory LPS^*24*^. A possible explanation for these findings may be related to the natural history of COVID-19 lung disease. Sequencing-based analysis of broncho-alveolar lavage fluid from patients hospitalized with COVID-19 lung disease has shown the presence of oropharyngeal flora, which are hypothesized to enter the lungs by aspiration^30^. The presence of organisms producing more inflammatory LPS in the oropharynx translocating to the lungs may potentiate inflammation during COVID-19 lung disease and lead to the need for respiratory support. Our findings support the hypothesis that a higher abundance of *Prevotella* and other species producing weakly immunogenic LPS corresponds to decreased abundance of more inflammatory LPS producing species. If aspiration and translocation occurs during COVID-19, the presence of organisms that produce less inflammatory LPS may limit inflammation in the lungs of COVID-19 patients.

### *Actinomyces* and Mycolic Acid Biosynthetic Pathway

A lower abundance of several *Actinomyces* were found to be predictive of the need for respiratory support in our RFC and an *Actinomyces* species was found as associated with the outcome via MaAsLiN2. *Actinomyces* are slow-growing, facultatively anaerobic, Gram-positive organisms and ubiquitous colonizers of the human body and environment^31,32^. Clinically, they are usually associated with slow progressing infections of the head, neck, chest and pelvis^32^. They are likely a component of a healthy oropharyngeal microbiome, in a study of the oropharyngeal microbiome among healthy adults, higher *Actinomyces* abundance was associated with decreased systemic inflammation^33^. They also are capable of biosynthesis of a wide variety of biologically active compounds including mycolic acid^34^. A lower abundance of the pathway for mycolic acid biosynthesis was a top predictor of the need for respiratory support in our RFC model and was also associated with the outcome by MaAsLiN2. *Actinomyces* is the only genera found to effect COVID-19 in this study hypothesized to be capable of mycolic acid production. An anti-inflammatory effect, possibly via mycolic acid biosynthesis, may be why a higher abundance of these organisms and this metabolic pathway is predictive of not requiring respiratory support.

### The Potential Protective Effect of Commensals

The predominant effect that we observed was that a decrease in the abundance of several commensal organisms and an increased abundance of bacterial products synthesis pathways of the oropharyngeal microbiome is the primary predictor of the need for respiratory support in COVID-19. The finding that the bacteria of the oropharyngeal microbiome are potentially protective against severe COVID-19 fits with observational data about the treatment of COVID-19 patients with antibiotics. These studies suggest that treatment of COVID-19 with antibiotics does not reduce mortality and that secondary bacterial infection is uncommon^35,36^. Our findings run counter to the hypothesis that the oropharynx is primarily a source of opportunistic pathogens that gain access to the lungs during the course of COVID-19^30^.

If the predominant effect were that the presence of harmful or pathogenic bacteria in the oropharyngeal microbiome contributing to severe COVID-19, one might expect treatment with antibiotics to be beneficial. Our findings are more consistent with the results of animal-model experiments with influenza, that suggest that treatment with antibiotics is potentially harmful due to their effect on beneficial commensal organisms. In mice challenged with influenza who had normal upper airway microbiomes, macrophages activated genes associated with anti-viral activity such as interferon-gamma, while those who were treated with antibiotics failed to activate these pathways and had more severe lung disease^9^. In another study, antibiotic treatment prior to influenza challenge impaired dendritic cell priming and migration to draining lymph nodes that ultimately led to impaired development of T-cell mediated adaptive immunity^37^. In COVID-19, the oropharyngeal microbiome may play a similar role, aiding the development of an effective anti-viral response that limits severe disease manifestations. In this context, the microbiome was demonstrated to be critical to an effective immune response to viral infection^8,9^.

### Strengths and Limitations

Our strengths include our enrollment of patients within the Emergency Department during acute presentation of the disease, prospective data collection, use of metagenomic sequencing, and use of two independent analysis techniques to verify our results. The enrollment and collection of samples within the Emergency Department has allowed us to sample the microbiome of patients early in disease course before medical intervention. We excluded any patients with self-reported symptoms longer than 14 days at time of collection to focus our analysis on the acute phase of the COVID-19. Our characterization of the oropharyngeal microbiome shows us features that can be predictive of disease course and potentially a target for therapeutics. In addition, the use of metagenomic sequencing for microbiome characterization has enabled us to determine what bacterial metabolic pathways could potentially affect disease course as opposed to just genus-level information provided by 16S rRNA sequencing. Although some microbiome features were also associated with age by MaAsLin2, these represent independent associations and would have been corrected for when determining associations with the need for respiratory support.

Weaknesses of this study include a single time-point in microbiome sampling from a single center and enrollment of a limiting number of patients presenting with acute COVID-19 early in the disease course. Single time-point sampling does not allow observation of how an individual oropharyngeal microbiome may change over the course of the disease. Although we enrolled 115 patients in the study, after focusing on the acute phase of COVID-19, only 50 COVID-19+ individuals with complete data were available for full analysis, which reduces statistical certainty. The reasons for incomplete data are multifactorial and include difficulties conducting clinical research during the COVID-19 pandemic. We developed a method to limit research staff contact with patients to prevent the spread of COVID-19 by having nursing staff collect specimens during routine clinical care after verbal consent. Although we successfully protected our staff, this necessitated the need for follow up to collect information on symptoms and symptom duration, which is challenging among an Emergency Department population, and led to missing clinical data and later withdrawal of consent.

## Conclusions

We demonstrate a relationship between disease manifestations of COVID-19 and the oropharyngeal microbiome. Specifically, the decreased abundance of some organisms, primarily *P. salivae*, is predictive of patients requiring respiratory support. We show that the presence of metabolic pathways for bacterial products such as LPS and mycolic acid are also predictive of not requiring respiratory support, implying that the presence of bacteria producing these products has a positive impact on disease course. Together, these findings suggest that the presence of beneficial commensal bacteria in the upper airway has the potential to prevent or mitigate pulmonary manifestations of COVID-19. Thus, our study underscores that the interaction between the oropharyngeal microbiome and respiratory viruses such as SARS-CoV2 could potentially be harnessed for diagnostic and therapeutic purposes.

## Methods

### Enrollment

Patients presenting with COVID-19 symptoms at the UMass Memorial Medical Center Emergency Department or while admitted to UMass Memorial COVID-19 treatment units were approached for enrollment in the study. Some individuals had known COVID-19 status when approached on inpatient COVID-19 wards, but the majority were approached in the Emergency Department prior to receiving results of clinical testing. Enrollment and sample collection took place April 2020 through March 2021, this occurred before vaccines were widely available and no subjects had been vaccinated against COVID-19. Enrolled patients were followed prospectively through the Electronic Medical Record (EMR). We collected information on disease outcomes of COVID-19 for their initial visit including need for respiratory support, the results of clinical laboratory testing, and mortality via the EMR. The Institutional Review Board at the University of Massachusetts Medical School approved this study (protocol # H00020145).

### Sample Collection and Processing

Oropharyngeal samples were collected using OMNIgene•ORAL collection kits (OMR-120, DNA Genotek). Briefly, the posterior oropharynx was swabbed for 30 seconds and collected as per manufacturer protocol. Samples were heated at 65-70°C for one hour^38^ to ensure SARS-CoV-2 inactivation and then stored frozen at -20°C. Upon thawing for nucleic acid extraction, samples were treated with 5ul Proteinase K (P8107S, New England Biolabs) for 2 hours at 50°C, then extracted using ZymoBIOMICS DNA/RNA Miniprep Kits (R2002, Zymo Research) as per manufacture protocol. DNA sequencing libraries were constructed using the Nextera XT DNA Library Prep Kit (FC-131-1096, Illumina) and sequenced on a NextSeq 500 Sequencing System as 2 × 150 nucleotide paired-end reads.

### Classification of Samples

Samples were classified as being collected from a patient with acute COVID-19 (COVID+) if they had a documented clinical testing that was positive rtPCR testing for SARS-CoV2 and self-reported symptoms for 14 days or less. The need for respiratory support was classified as positive if the patient required any intervention to support breathing. This included supplemental oxygen via nasal cannula or face mask, non-invasive possible pressure ventilation, or intubation. If a patient had a Do Not Intubate (DNI) order but went on to die of COVID-19 symptoms, we considered that patient has having respiratory failure severe enough to require intubation and classified the sample as being from a patient who was intubated. Patients were considered as having in-hospital mortality from COVID-19 if this was listed as a cause of death on hospital death records.

### Sequence Processing and Analysis

Shotgun metagenomic reads were first trimmed and quality filtered to remove sequencing adapters and host contamination using Trimmomatic^39^ and Bowtie2^40^, respectively, as part of the KneadData pipeline version 0.7.2 (https://huttenhower.sph.harvard.edu/kneaddata/). As in our previous work^41,42^, reads were then profiled for microbial taxonomic abundances and metabolic pathways using Metaphlan3 and HUMAnN3, respectively^43^ (https://www.biorxiv.org/content/10.1101/2020.11.19.388223v1).

### Microbiome-clinical factors modeling

To determine the association between bacterial species abundance and COVID-19 diagnosis, we performed a non-parametric Wilcoxon Rank Sum test for species with at least 5% prevalence and a minimal average relative abundance of 0.01% across all samples (n=115; 74 COVID-19+ and 41 COVID-19–) with the Bonferroni correction for multiple comparisons. To identify oropharyngeal bacteria and clinical covariates that are predictive of respiratory support in COVID-19+ patients and compare their relative contributions, we developed and ran a Random Forest Classification (RFC)-based pipeline in R. For each subset of data, the pipeline was run six times from six different random seeds and statistics for the model’s classification performance and variables contribution to class discrimination were calculated for each seed. The first step of the pipeline is a leave-one-out cross-validation split of the data. The resulting train set is then used for the following steps of the pipeline. Feature selection using Boruta^44^ is then run in a leave-one-out cross-validation scheme to select a subset of variables that are discriminatory. The Boruta-selected variables were then used to train a RFC, using the ranger package^14^. The resulting RFC model was then used to predict the left-out sample. Thus, the performance of our model is calculated based on the aggregated predictions of left-out data. The top 18 most important variables were then used to run MaAsLin2^15^ to examine their multivariate association. The FDR corrected p-value and coefficient are shown on the violin plots. Plots were generated in R using the ggplot2 package^45^ and color palettes from the calecopal package (https://github.com/an-bui/calecopal).

## Supporting information

Species Origin of Metabolic Pathways

RFC on Bacterial Species Statistics

RFC on Metabolic Pathways Statistics

## Data Availability

All data produced in the present study are available upon reasonable request to the authors

## ACKNOWLEDGEMENTS

We would like to thank the UMass Memorial Medical Center Emergency Department Staff, especially the nursing and resident physicians for making it possible to collect biological samples from COVID-19 patients with acute and sometimes severe symptoms within in the Emergency Department. Thank you to the Human Patients Institutional Review Board at the University of Massachusetts Medical School and especially A. Blodgett for their guidance in helping to design and implement the human patient protocol early in the pandemic. Thank you to The Society for Academic Emergency Medicine as well as the Dean of University of Massachusetts Medical School and the many donors who through their financial support made this work possible. We would also like to thank the NIH for funding that provided salary support for this work (1RF1AG067483-01).

## AUTHOR CONTRIBUTIONS

ESB, JPH, BAM, and AM, conceived and led the study. JPH, ESB, CT supervised the conduct of the study and data collection. LC, MMS, SM, CT, and PD managed the clinical data, including quality control. LC and MMS handled the sample collection and storage. DW managed sample extraction and sequencing, and performed metagenomic profiling. ALZ and VB provided statistical advice on study design and performed all ML modeling and microbiome-clinical covariates-clinical outcome statistical analysis. ESB, ALZ, VB and JPH wrote the manuscript with input from all authors.

## Figure Legends

**Figure S1 Bacterial Genus Origin of Detected Metabolic Pathways Predicitive of Need For Respiratory Support**. Panel A, Contribution of detected bacterial genera to pathway abundance of CMP-3-deoxy-D-manno-octusonate in patiens who did and did not go on to require respiratory support. Panel B, Contribution of detected bacterial genera to pathway abundance of Lipid IV A biosynthesis in patients who did and did not go on to require respiratory support. Noteable is the presence of *Pseudomonas* contributing to the detected CMP-3-deoxy-D-manno-octusonate pathway abundance and increased abundance of *Aggrigatibacter* contributing to the Lipid IV A pathway.

## References

1. Dong, E., Du, H. & Gardner, L. An interactive web-based dashboard to track COVID-19 in real time. The Lancet Infectious Diseases 20, 533–534 (2020).

2. Bai, Y., et al. Presumed Asymptomatic Carrier Transmission of COVID-19. JAMA (2020).

3. Gallo Marin, B., et al. Predictors of COVID-19 severity: A literature review. Rev Med Virol 31, 1–10 (2021).

4. Covino, M., et al. Predicting In-Hospital Mortality in COVID-19 Older Patients with Specifically Developed Scores. J Am Geriatr Soc 69, 37–43 (2021).

5. Baker, K.F., et al. National Early Warning Score 2 (NEWS2) to identify inpatient COVID-19 deterioration: a retrospective analysis. Clin Med (Lond) 21, 84–89 (2021).

6. Beck, D.B. & Aksentijevich, I. Susceptibility to severe COVID-19. Science 370, 404–405 (2020).

7. Man, W.H., de Steenhuijsen Piters, W.A. & Bogaert, D. The microbiota of the respiratory tract: gatekeeper to respiratory health. Nat Rev Microbiol 15, 259–270 (2017).

8. Short, K.R., et al. Bacterial lipopolysaccharide inhibits influenza virus infection of human macrophages and the consequent induction of CD8+ T cell immunity. J Innate Immun 6, 129–139 (2014).

9. Abt, M.C., et al. Commensal bacteria calibrate the activation threshold of innate antiviral immunity. Immunity 37, 158–170 (2012).

10. McCullers, J.A. The co-pathogenesis of influenza viruses with bacteria in the lung. Nat Rev Microbiol 12, 252–262 (2014).

11. Avadhanula, V., et al. Respiratory viruses augment the adhesion of bacterial pathogens to respiratory epithelium in a viral species-and cell type-dependent manner. J Virol 80, 1629–1636 (2006).

12. Haran, J.P., et al. Alzheimer’s Disease Microbiome Is Associated with Dysregulation of the Anti-Inflammatory P-Glycoprotein Pathway. mBio 10(2019).

13. Wipperman, M.F., et al. Gastrointestinal microbiota composition predicts peripheral inflammatory state during treatment of human tuberculosis. Nat Commun 12, 1141 (2021).

14. Wright, M.N. & Ziegler, A. ranger: A Fast Implementation of Random Forests for High Dimensional Data in C++ and R. 2017 77, 17 (2017).

15. Mallick, H., et al. (2021).

16. Franzosa, E.A., et al. Species-level functional profiling of metagenomes and metatranscriptomes. Nat Methods 15, 962–968 (2018).

17. Norskov-Lauritsen, N. Classification, identification, and clinical significance of Haemophilus and Aggregatibacter species with host specificity for humans. Clin Microbiol Rev 27, 214–240 (2014).

18. Ventero, M.P., et al. Nasopharyngeal Microbial Communities of Patients Infected With SARS-CoV-2 That Developed COVID-19. Front Microbiol 12, 637430 (2021).

19. Yang, F., et al. Saliva microbiomes distinguish caries-active from healthy human populations. ISME J 6, 1–10 (2012).

20. Chakraborty, S. The 2019 Wuhan Outbreak Could be Caused by the Bacteria Prevotella, Which is Aided by the Coronavirus-Prevotella is Present (Sometimes in Huge Amounts) in Patients from Two Studies in China and One in Hong Kong. OSF [Preprint]. doi 10(2020).

21. Khan, A.A. & Khan, Z. COVID-2019-associated overexpressed Prevotella proteins mediated hostpathogen interactions and their role in coronavirus outbreak. Bioinformatics 36, 4065–4069 (2020).

22. Larsen, J.M. The immune response to Prevotella bacteria in chronic inflammatory disease. Immunology 151, 363–374 (2017).

23. Khatiwada, S. & Subedi, A. Lung microbiome and coronavirus disease 2019 (COVID-19): Possible link and implications. Hum Microb J 17, 100073 (2020).

24. Larsen, J.M., et al. Chronic obstructive pulmonary disease and asthma-associated Proteobacteria, but not commensal Prevotella spp., promote Toll-like receptor 2-independent lung inflammation and pathology. Immunology 144, 333–342 (2015).

25. Brix, S., Eriksen, C., Larsen, J.M. & Bisgaard, H. Metagenomic heterogeneity explains dual immune effects of endotoxins. J Allergy Clin Immunol 135, 277–280 (2015).

26. Goldman, R., Doran, C., Kadam, S. & Capobianco, J. Lipid A precursor from Pseudomonas aeruginosa is completely acylated prior to addition of 3-deoxy-D-manno-octulosonate. Journal of Biological Chemistry 263, 5217–5223 (1988).

27. Brozek, K.A. & Raetz, C. Biosynthesis of lipid A in Escherichia coli. Acyl carrier protein-dependent incorporation of laurate and myristate. Journal of Biological Chemistry 265, 15410–15417 (1990).

28. Beutler, B. & Poltorak, A. The sole gateway to endotoxin response: how LPS was identified as Tlr4, and its role in innate immunity. Drug Metabolism and Disposition 29, 474–478 (2001).

29. Goldberg, J.B. & Pler, G. Pseudomonas aeruginosa lipopolysaccharides and pathogenesis. Trends in microbiology 4, 490–494 (1996).

30. Bao, L., et al. Oral Microbiome and SARS-CoV-2: Beware of Lung Co-infection. Front Microbiol 11, 1840 (2020).

31. Bowden, G.H.W. Actinomyces, Propionibacterium propionicus, and Streptomyces. in Medical Microbiology (ed. Baron, S.) (University of Texas Medical Branch at Galveston Copyright © 1996, The University of Texas Medical Branch at Galveston., Galveston (TX), 1996).

32. Kononen, E. & Wade, W.G. Actinomyces and related organisms in human infections. Clin Microbiol Rev 28, 419–442 (2015).

33. Demmer, R.T., et al. The subgingival microbiome, systemic inflammation and insulin resistance: The Oral Infections, Glucose Intolerance and Insulin Resistance Study. J Clin Periodontol 44, 255–265 (2017).

34. Collins, M., Goodfellow, M., Minnikin, D. & Alderson, G. Menaquinone composition of mycolic acid-containing actinomycetes and some sporoactinomycetes. Journal of applied bacteriology 58, 77–86 (1985).

35. Chedid, M., et al. Antibiotics in treatment of COVID-19 complications: a review of frequency, indications, and efficacy. J Infect Public Health 14, 570–576 (2021).

36. Langford, B.J., et al. Antibiotic prescribing in patients with COVID-19: rapid review and meta-analysis. Clin Microbiol Infect 27, 520–531 (2021).

37. Ichinohe, T., et al. Microbiota regulates immune defense against respiratory tract influenza A virus infection. Proc Natl Acad Sci U S A 108, 5354–5359 (2011).

38. Rabenau, H.F., et al. Stability and inactivation of SARS coronavirus. Med Microbiol Immunol 194, 1–6 (2005).

39. Bolger, A.M., Lohse, M. & Usadel, B. Trimmomatic: a flexible trimmer for Illumina sequence data. Bioinformatics 30, 2114–2120 (2014).

40. Langmead, B. & Salzberg, S.L. Fast gapped-read alignment with Bowtie 2. Nat Methods 9, 1–3 (2012).

41. Haran, J.P., et al. Alzheimer’s Disease Microbiome Is Associated with Dysregulation of the Anti-Inflammatory P-Glycoprotein Pathway. mBio 10(2019).

42. Haran, J.P., Bucci, V., Dutta, P., Ward, D. & McCormick, B. The nursing home elder microbiome stability and associations with age, frailty, nutrition, and physical location. Journal of medical microbiology 67, 40–51 (2018).

43. Truong, D.T., et al. MetaPhlAn2 for enhanced metagenomic taxonomic profiling. Nat Methods 12(2015).

44. Kursa, M.B. & Rudnicki, W.R. Feature Selection with the Boruta Package. 2010 36, 13 (2010).

45. Wickham, H. ggplot2: Elegant Graphics for Data Analysis, (Springer-Verlag, New York, 2016).

