## Supplementary figures and images for "Oropharyngeal Microbiome Profiled at Admission is Predictive of the Need for Respiratory Support Among COVID-19 Patients"

### Species Origin of Metabolic Pathways

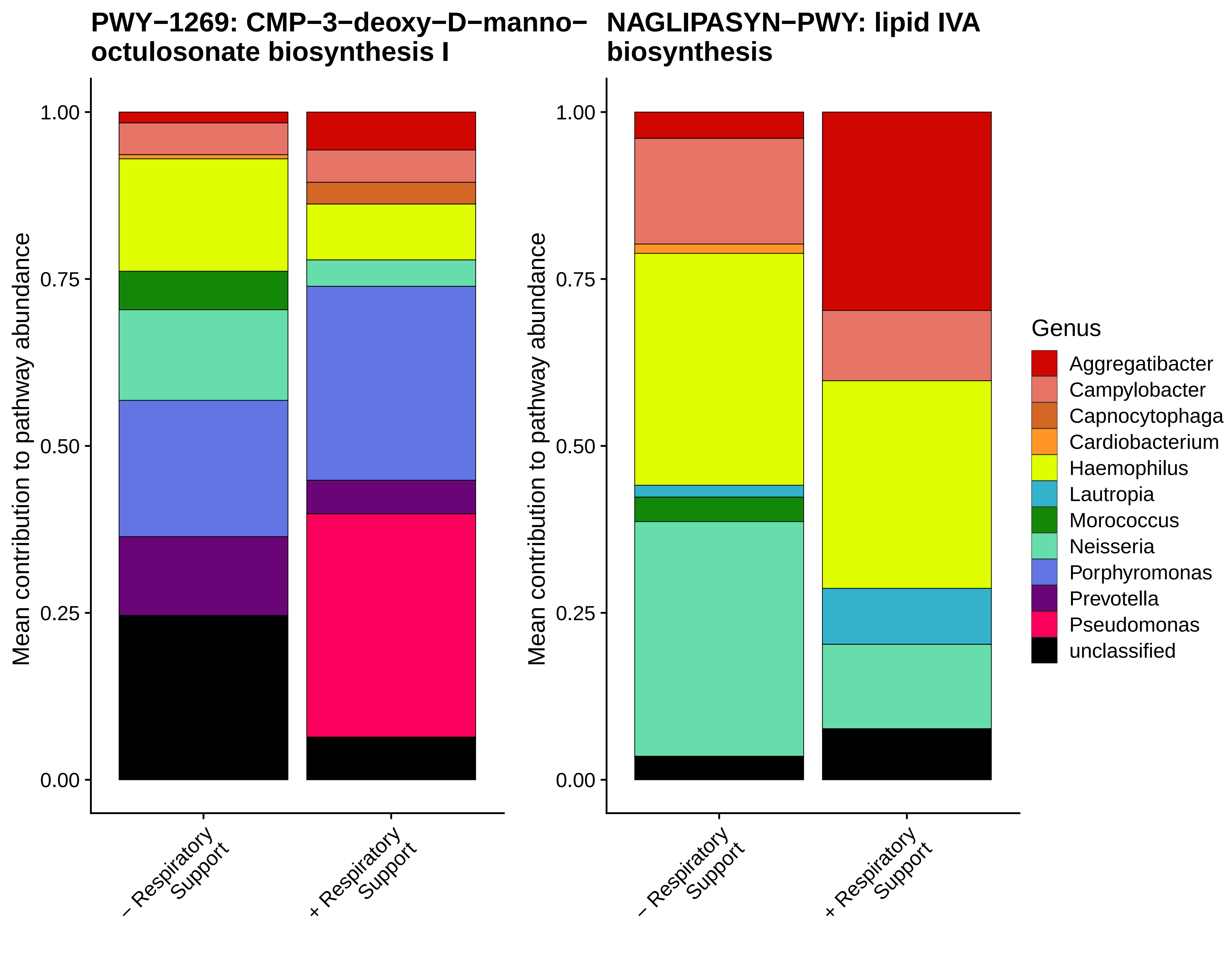
