## Supplementary material for "Oropharyngeal Microbiome Profiled at Admission is Predictive of the Need for Respiratory Support Among COVID-19 Patients": RFC on Bacterial Species Statistics

| Model | Seed | TP | FP | TN | FN | Sensitivity | Specificity | Precision | Accuracy | F1 |
|---|---|---|---|---|---|---|---|---|---|---|
| Bacterial Abundance | 111 | 35 | 11 | 1 | 3 | 0.92 | 0.08 | 0.76 | 0.72 | 0.83 |
| Bacterial Abundance | 112 | 35 | 11 | 1 | 3 | 0.92 | 0.08 | 0.76 | 0.72 | 0.83 |
| Bacterial Abundance | 113 | 35 | 11 | 1 | 3 | 0.92 | 0.08 | 0.76 | 0.72 | 0.83 |
| Bacterial Abundance | 114 | 35 | 11 | 1 | 3 | 0.92 | 0.08 | 0.76 | 0.72 | 0.83 |
| Bacterial Abundance | 115 | 35 | 10 | 2 | 3 | 0.92 | 0.17 | 0.78 | 0.74 | 0.84 |
| Bacterial Abundance | 116 | 35 | 10 | 2 | 3 | 0.92 | 0.17 | 0.78 | 0.74 | 0.84 |
| CC + Shannon Diversity | 111 | 36 | 10 | 2 | 2 | 0.95 | 0.17 | 0.78 | 0.76 | 0.86 |
| CC + Shannon Diversity | 112 | 36 | 10 | 2 | 2 | 0.95 | 0.17 | 0.78 | 0.76 | 0.86 |
| CC + Shannon Diversity | 113 | 36 | 10 | 2 | 2 | 0.95 | 0.17 | 0.78 | 0.76 | 0.86 |
| CC + Shannon Diversity | 114 | 36 | 10 | 2 | 2 | 0.95 | 0.17 | 0.78 | 0.76 | 0.86 |
| CC + Shannon Diversity | 115 | 36 | 10 | 2 | 2 | 0.95 | 0.17 | 0.78 | 0.76 | 0.86 |
| CC + Shannon Diversity | 116 | 36 | 10 | 2 | 2 | 0.95 | 0.17 | 0.78 | 0.76 | 0.86 |
| CC + Bacterial Abundance | 111 | 35 | 9 | 3 | 3 | 0.92 | 0.25 | 0.80 | 0.76 | 0.85 |
| CC + Bacterial Abundance | 112 | 36 | 10 | 2 | 2 | 0.95 | 0.17 | 0.78 | 0.76 | 0.86 |
| CC + Bacterial Abundance | 113 | 36 | 11 | 1 | 2 | 0.95 | 0.08 | 0.77 | 0.74 | 0.85 |
| CC + Bacterial Abundance | 114 | 36 | 9 | 3 | 2 | 0.95 | 0.25 | 0.80 | 0.78 | 0.87 |
| CC + Bacterial Abundance | 115 | 36 | 10 | 2 | 2 | 0.95 | 0.17 | 0.78 | 0.76 | 0.86 |
| CC + Bacterial Abundance | 116 | 35 | 10 | 2 | 3 | 0.92 | 0.17 | 0.78 | 0.74 | 0.84 |
| CC + Bacterial Abund. + Shannon Div. | 111 | 36 | 10 | 2 | 2 | 0.95 | 0.17 | 0.78 | 0.76 | 0.86 |
| CC + Bacterial Abund. + Shannon Div. | 112 | 36 | 9 | 3 | 2 | 0.95 | 0.25 | 0.80 | 0.78 | 0.87 |
| CC + Bacterial Abund. + Shannon Div. | 113 | 36 | 9 | 3 | 2 | 0.95 | 0.25 | 0.80 | 0.78 | 0.87 |
| CC + Bacterial Abund. + Shannon Div. | 114 | 36 | 10 | 2 | 2 | 0.95 | 0.17 | 0.78 | 0.76 | 0.86 |
| CC + Bacterial Abund. + Shannon Div. | 115 | 35 | 9 | 3 | 3 | 0.92 | 0.25 | 0.80 | 0.76 | 0.85 |
| CC + Bacterial Abund. + Shannon Div. | 116 | 35 | 10 | 2 | 3 | 0.92 | 0.17 | 0.78 | 0.74 | 0.84 |
