## Supplementary material for "Oropharyngeal Microbiome Profiled at Admission is Predictive of the Need for Respiratory Support Among COVID-19 Patients": RFC on Metabolic Pathways Statistics

| Model | seed | TP | FP | TN | FN | sensitivity | specificity | precision | accuracy | F1 |
|---|---|---|---|---|---|---|---|---|---|---|
| Pathway Abundance | 111.00 | 33.00 | 11.00 | 1.00 | 5.00 | 0.87 | 0.08 | 0.75 | 0.68 | 0.80 |
| Pathway Abundance | 112.00 | 34.00 | 12.00 | 0.00 | 4.00 | 0.89 | 0.00 | 0.74 | 0.68 | 0.81 |
| Pathway Abundance | 113.00 | 34.00 | 12.00 | 0.00 | 4.00 | 0.89 | 0.00 | 0.74 | 0.68 | 0.81 |
| Pathway Abundance | 114.00 | 33.00 | 11.00 | 1.00 | 5.00 | 0.87 | 0.08 | 0.75 | 0.68 | 0.80 |
| Pathway Abundance | 115.00 | 32.00 | 12.00 | 0.00 | 6.00 | 0.84 | 0.00 | 0.73 | 0.64 | 0.78 |
| Pathway Abundance | 116.00 | 33.00 | 11.00 | 1.00 | 5.00 | 0.87 | 0.08 | 0.75 | 0.68 | 0.80 |
| Pathway Abund. + CC | 111.00 | 34.00 | 11.00 | 1.00 | 4.00 | 0.89 | 0.08 | 0.76 | 0.70 | 0.82 |
| Pathway Abund. + CC | 112.00 | 35.00 | 10.00 | 2.00 | 3.00 | 0.92 | 0.17 | 0.78 | 0.74 | 0.84 |
| Pathway Abund. + CC | 113.00 | 34.00 | 11.00 | 1.00 | 4.00 | 0.89 | 0.08 | 0.76 | 0.70 | 0.82 |
| Pathway Abund. + CC | 114.00 | 34.00 | 10.00 | 2.00 | 4.00 | 0.89 | 0.17 | 0.77 | 0.72 | 0.83 |
| Pathway Abund. + CC | 115.00 | 34.00 | 11.00 | 1.00 | 4.00 | 0.89 | 0.08 | 0.76 | 0.70 | 0.82 |
| Pathway Abund. + CC | 116.00 | 34.00 | 10.00 | 2.00 | 4.00 | 0.89 | 0.17 | 0.77 | 0.72 | 0.83 |
